# Cardiopulmonary imaging utilization and findings among hospitalized COVID-19 patients in Latin America (From RIMAC: Registry IMAging Cardiopulmonary among hospitalized COVID-19 patients in LATAM)

**DOI:** 10.1101/2022.01.10.22269002

**Authors:** Salvador V Spina, Marcelo L Campos Vieira, César J. Herrera, Ana G. Múnera Echeverri, Pamela Rojo, Alma S Arrioja Salazar, Zuilma Y Vázquez Ortiz, Roberto Baltodano Arellano, Graciela Reyes, Rocío Aceves Millán, Juan A Calderón González, Ana C Camarozano, Edgar Avilés, Marco A Cabrera, María F Grande Ratti, Jorge Lowenstein, Rodrigo Hernández Vyhmeister, Pamela Piña Santana, Jaime A. Ibarra Burgos, Alejandra Rivera, Beatriz A Fernández Campos, Kelly M Cupe Chacalcaje, Mariela De Santos, Tania R Afonso, Tomás Miranda Aquino, Ana L Lalyre Acosta, Beatriz Domínguez, Federico Campos, Sergio M Alday Ramirez, Angela V Cachicatari Beltran, Daniela Alvarez, Patricia de Oliveira Roveri, Carlos Rosales Ixcamparij, Ender López, Pedro Vargas, Maximiliano Flores Flamand, Rosa L López Martínez, Luciana Meza, Samira Saady Morthy, Rudy Ovalle, Stalin Martínez, Oscar A Pérez Orpinel, Mauricio Potito, Otto Orellana, Jorge Marte Baez, Consuelo Orihuela Sandoval, Marcos Granillo Fernandez, Rohit Loomba, Saúl Flores, José M Hernández Hernández, Ricardo Pignatelli

## Abstract

**Objectives:** To describe the use and findings of cardiopulmonary imaging - chest X-ray (cX-ray), echocardiography (cEcho), chest CT (cCT), lung ultrasound (LUS)) and/or cardiac magnetic resonance imaging (cMRI) - in COVID-19-associated hospitalizations in Latin America (LATAM)

**Background:** The SARS-Cov-2 is one of the largest and most active threats to healthcare in living memory. There is an information gap on imaging services resources (ISR) used and their findings during the pandemic in LATAM.

**Methods:** This was a multicenter, prospective, observational study of COVID-19 inpatients conducted from March to December 2020 from 12 high-complexity centers in nine LATAM countries. Adults (> 18 yrs) with at least one imaging modality performed, followed from admission until discharge and/or in-hospital death, were included.

**Results:** We studied 1435 hospitalized patients (64% males) with a median age of 58 years classified into three regions: 262 from Mexico (Mx), 428 from Central America and Caribbean (CAC), and 745 from South America (SAm). More frequent comorbidities were overweight/obesity (61%), hypertension (45%), and diabetes (27%). During hospitalization, 58% were admitted to ICU. The in-hospital mortality was 28% (95%CI 25-30) highest in Mx (37%).

The most frequent cardiopulmonary imaging performed were cCT (61%)-more frequent in Mx and SAm-, and cX-ray (46%) -significantly used in CAC-. The cEcho was carried out in 18%, similarly among regions, and LUS in 7%, more frequently in Mx. The cMRI was performed in only one patient in the cohort. Abnormal findings on the cX-ray were related to peripheral (63%) or basal infiltrates (52%), and in cCT with ground glass infiltrates (89%). Both were more commonly in Mx. In LUS, interstitial syndrome (56%) was the most related abnormal finding, predominantly in Mx and CAC.

**Conclusions:** The use and findings of cardiopulmonary imaging in LATAM varied between regions and may have been influenced by clinical needs, the personnel protection measures and/or hospitalization location.

**Condensed Abstract:** The SARS-Cov-2 is one of the largest and most active threats to healthcare in living memory. There is limited information on imaging services resources (ISR) used and their findings during the pandemic in LATAM.

To our knowledge, RIMAC aimed the first international, multicenter study at registering the use and findings of cardiopulmonary imaging modalities performed for the diagnosis, prognosis, and treatment of patients hospitalized for infection with SARS-CoV-2 in Latin America. We studied their demographic parameters, comorbidities, in-hospital events, laboratory results, and treatments focusing on their impact in clinical complications.

## Introduction

The COVID-19 pandemic is one of the largest and most active threats to healthcare in living memory. As the impact of the virus continues, systems of care around the world have responded with unprecedented protective measures.

SARS-Cov-2 predominantly affects adults, and disease severity increases with age and number of comorbidities. COVID-19 infection is mainly characterized by upper airway inflammation, which can progress into interstitial pneumonia and eventually to acute respiratory distress syndrome (ARDS) in the most severe cases. Cardiovascular complications such as thromboembolic phenomena, acute coronary syndrome, heart failure, as well as renal dysfunction are known to occur (1-4).

Cardiopulmonary imaging plays an essential role in the diagnosis of SARS-CoV-2 infection and its complications. Imaging can assess the extent of disease, prognosis, and evaluation of therapeutic interventions (1-7). Imaging services resources (ISR) such as electrocardiogram (ECG) (8,9), chest X-ray (cX-ray) (10), echocardiogram (cEcho) (11-14), lung ultrasound (LUS) (15,16), and chest computed tomography (cCT) (17,18), have been at the front line of the pandemic. Each technique offers well-known advantages; however, despite the high number of cases of infection and deaths from COVID-19, their specific application and utilization in low and middle-income countries remains unknown.

Preventive distancing and biosecurity measures during testing can protect patients and staff, thus abbreviated evaluations and imaging interventions have become the norm. RIMAC aimed at registering the use and findings of cardiopulmonary imaging modalities performed for the diagnosis, prognosis, and treatment of patients hospitalized for infection with SARS-CoV-2. We studied their demographic parameters, comorbidities, in-hospital events, laboratory results, and concomitant treatments focusing on their impact in clinical complications.

## Methods

### Study design and patients

We conducted a multicenter, prospective, observational study that included adult patients with SARS-Cov-2 disease admitted from March to December 2020, in 12 high-complexity centers, level III or IV, from 9 countries -Argentina, Brazil, Colombia, Chile, Guatemala, Mexico, Panama, Peru, and the Dominican Republic-, whom were divided into three geographic regions.All the centers had availability of the total imaging modalities analyzed. The total number of referral beds in each center was greater than 185, which doubled and tripled according to the needs of each region. Inclusion criteria were age >18 years old, positive COVID-19 CRP, and/or COVID-19 positive IgM and IgG antibodies, hospitalized status, and at least one imaging modality performed according to each treating physician’s criteria (cX-ray, cEcho, LUS, cCT, or cMRI). The cohort included a consecutive sample of patients, followed up from admission until discharge and/or in-hospital death. Patients were prospectively monitored for major complications during hospitalization.

The exclusion criteria were: patients < 18 years old, lack of complete documentation on COVID-19 infection, non-hospitalized patients, failure to perform an imaging modality or inadequate quality for diagnosis and/or treatment (cX-ray,cEcho,LUS,cCT or cMRI).

### Data collection

Patient medical records were reviewed by the research team, and data (demographic, epidemiologial, clinical, imaging, laboratory, treatment and outcome) were retriev from electronic medical records using a standardized case report form (RIMAC Registry). Data was collected and confidentially stored in a database created for this purpose; each patient was assigned a de-identifying code. Local IRB approval from each center was obtained; researchers were not involved in direct care of the subjects. The obligation to have or not to have an informed consent was left to the discretion of each institution due to the nature of this study (prospective medical records review) with no intervention.

### Statistical analyses

All statistical analyses were conducted using STATA (version 17.0; StataCorp, College Station, TX). Descriptive variables are presented as numbers and percentages for categorical variables; mean (± standard deviation) for normally distributed continuous variables; and median (interquartile range) for non-normally distributed continuous variables.

We divided the cohort into three geographic regions for multiple comparisons. These were identified and agreed upon by the authors *a priori*: Mexico (Mx) as a representation of the Latin population of North America, Central America and Caribbean (CAC) that included Guatemala, Panamá, and the Dominican Republic; and South America (SAm) that included Argentina, Brazil, Chile, Colombia, and Perú.

Comparisons between regions of categorical variables were performed using Pearson’s χ2 test with Bonferroni’s correction. Meanwhile for continuous variables, the analysis of variance (ANOVA) or Kruskal−Wallis H test were used to compare the differences among the three groups as appropriate. In these cases of multiple comparison, only a value of p≤0.016 was considered statistically significant.

Additionally, univariate and multivariate logistic regression was performed to explore the association of factors that affect the likelihood of a specific clinical imaging modality being utilized. The specific imaging modality was the dependent variable (chest computed tomography, chest X-ray, echocardiogram, and lung ultrasound). The independent variables were clinical characteristics (comorbidities prior to admission) or variables related to complications (IOT, ICU admission). The statistical significance level was set at 0.05 (two-tailed), and odds ratios (OR) and their respective 95% confidence intervals (95%CI) were provided.

## Results

### Cohort overview

Of the 1549 patients initially recruited to the registry, 114 were excluded due to a lack of complete documentation on COVID-19 infection. There were 1435 hospitalized patients (64% males) included with a median age of 58 (SD 16.6) years: 262 from Mx, 428 from CAC, and 745 from SAm (Figure I). Figure II shows that the period of highest patient recruitment was from April to August 2020 in accordance with the first wave of COVID-19 in LATAM.

**Figure I:**
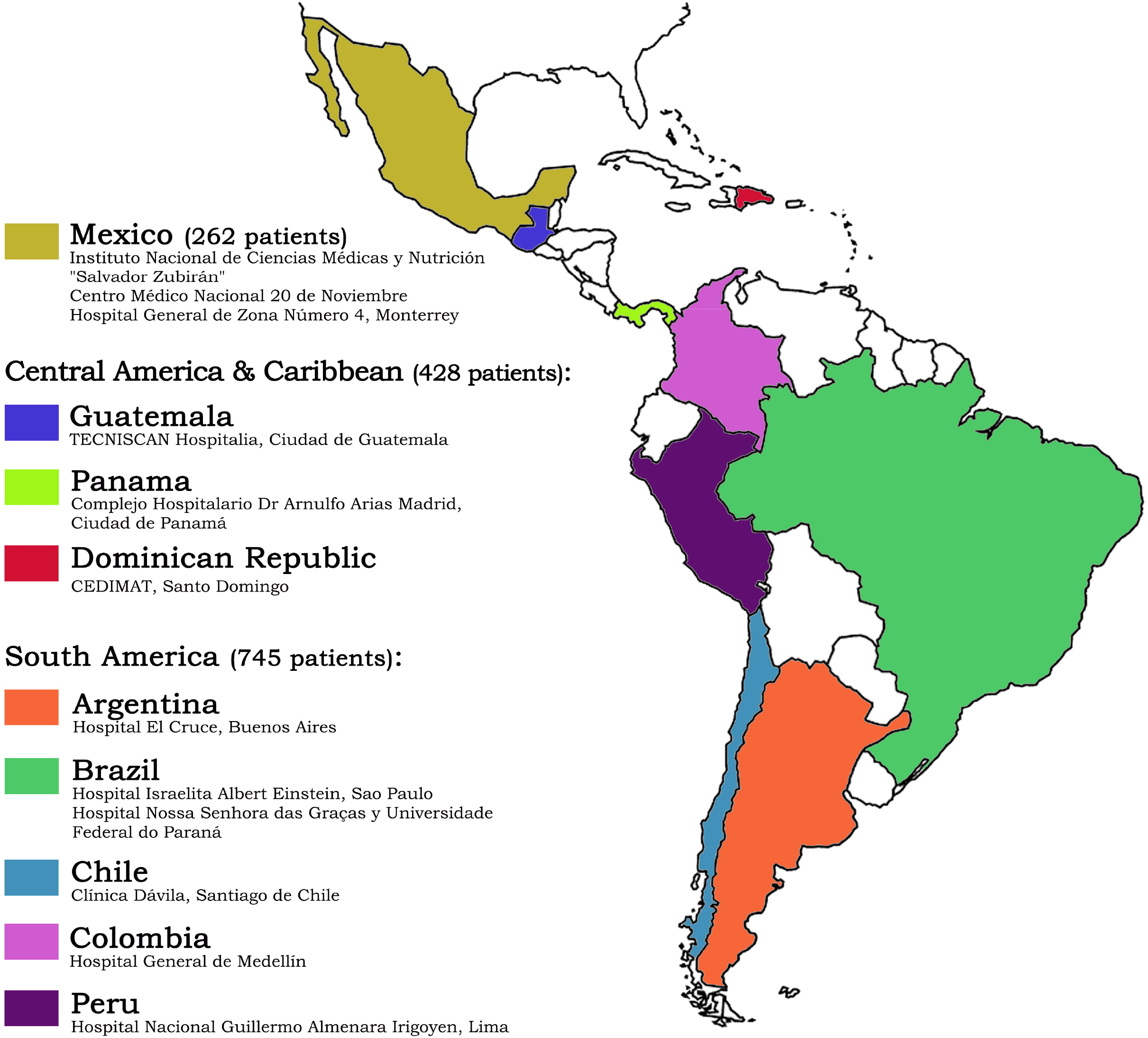
RIMAC Registry: Participating regions, countries and institutions (1435 patients)

**Figure II:**
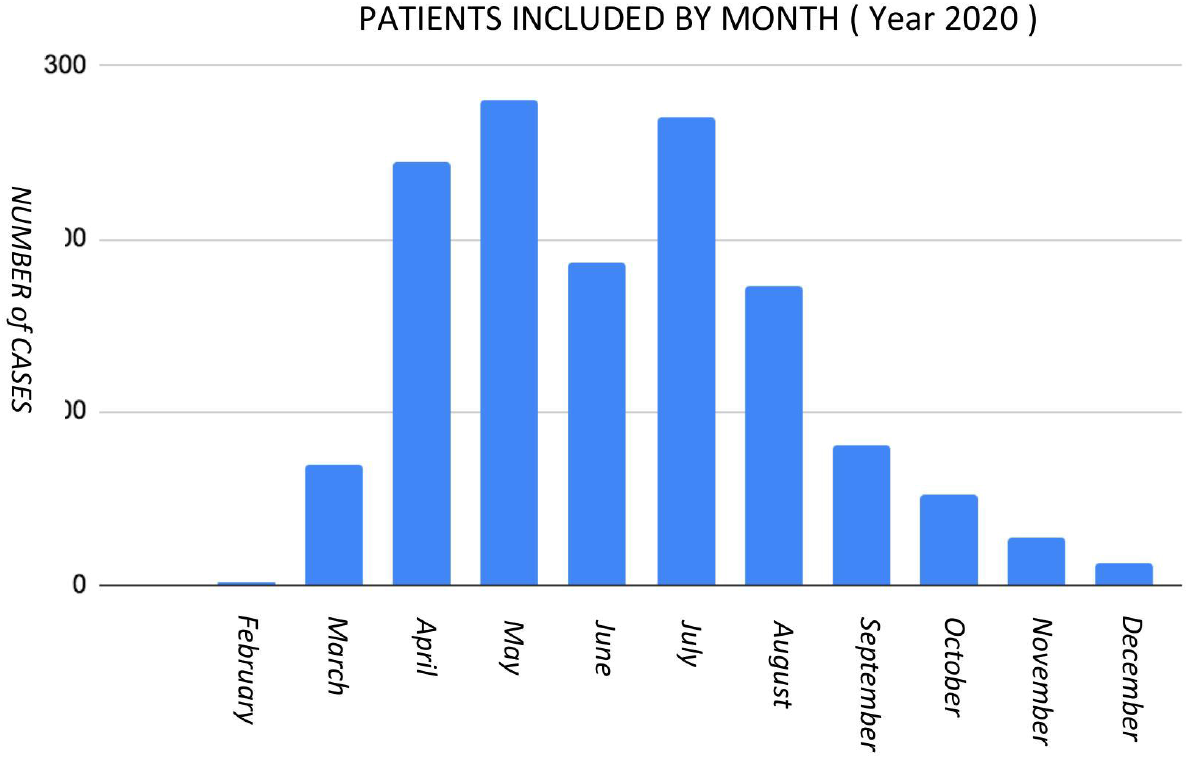
Patients included from February to December 2020 (1435)

A total of 708 patients (49%) were admitted to the ward: 727 (51%) patients were initially admitted to the ICU. This number subsequently increased to 836 (58%) during total hospital stay (most were Mexican patients) and this increase normalized the differences among regions. The median number of hospitalization days was 17 (SD 17.2), and the median ICU days was 13 (SD 14.2). This is significantly lower in CAC (Table 5).

The most common comorbidities were overweight/obesity (61%), hypertension (45%), and diabetes (27%). There were differences across regions: body mass index (BMI) was registered in 1180 patients, and overweight/obesity (BMI ≥25) was significantly more prevalent in Mexico especially due to obesity -BMI ≥30- (42%). Hypertension (HBP) was significantly higher in CAC (60%), and diabetes (DBT) predominated in Mx and CAC (34%). Ischemic heart disease, chronic obstructive pulmonary disease (COPD), renal failure, and myocardiopathy were below 7% of the population (Table 1).

**Table 1:**
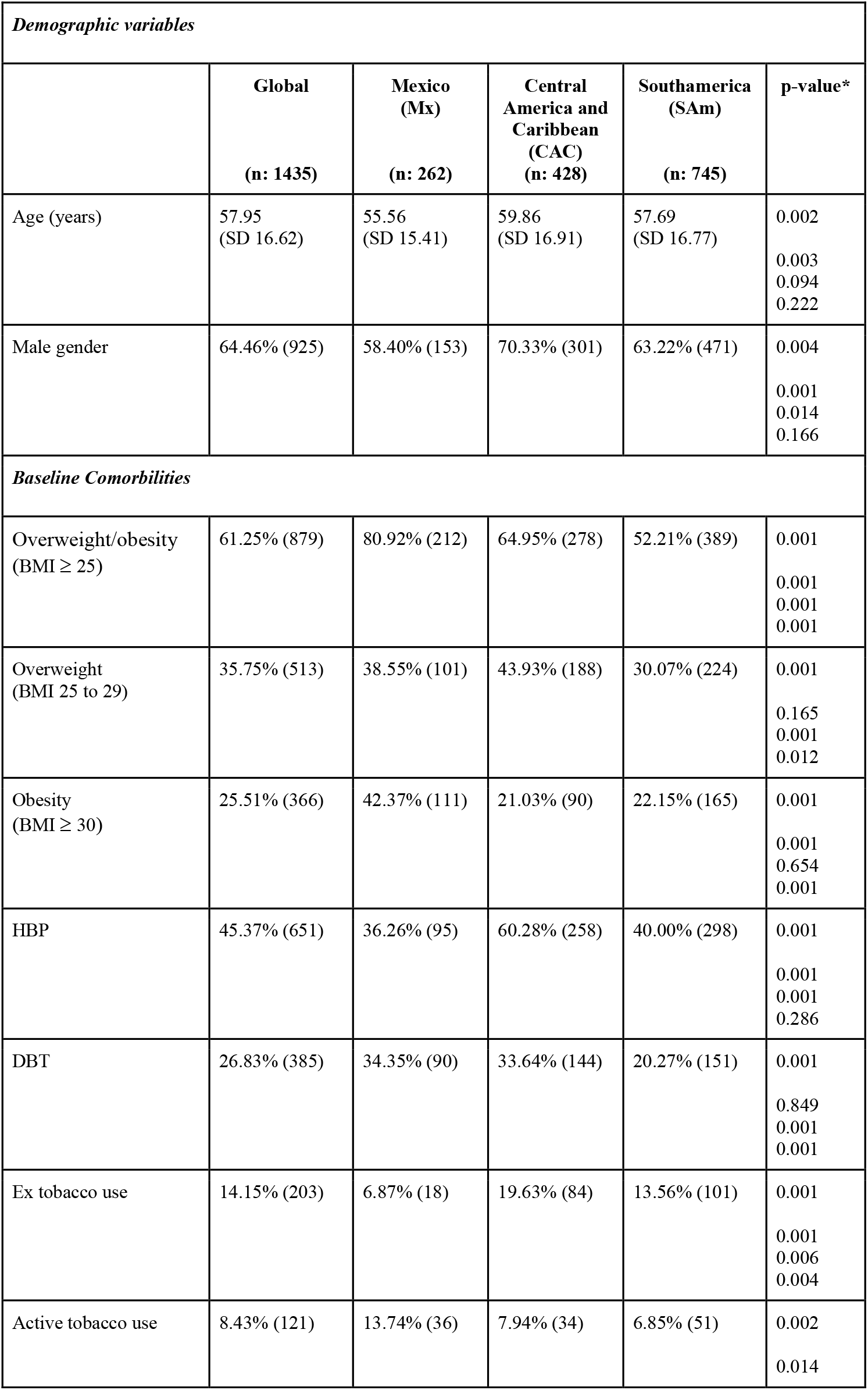
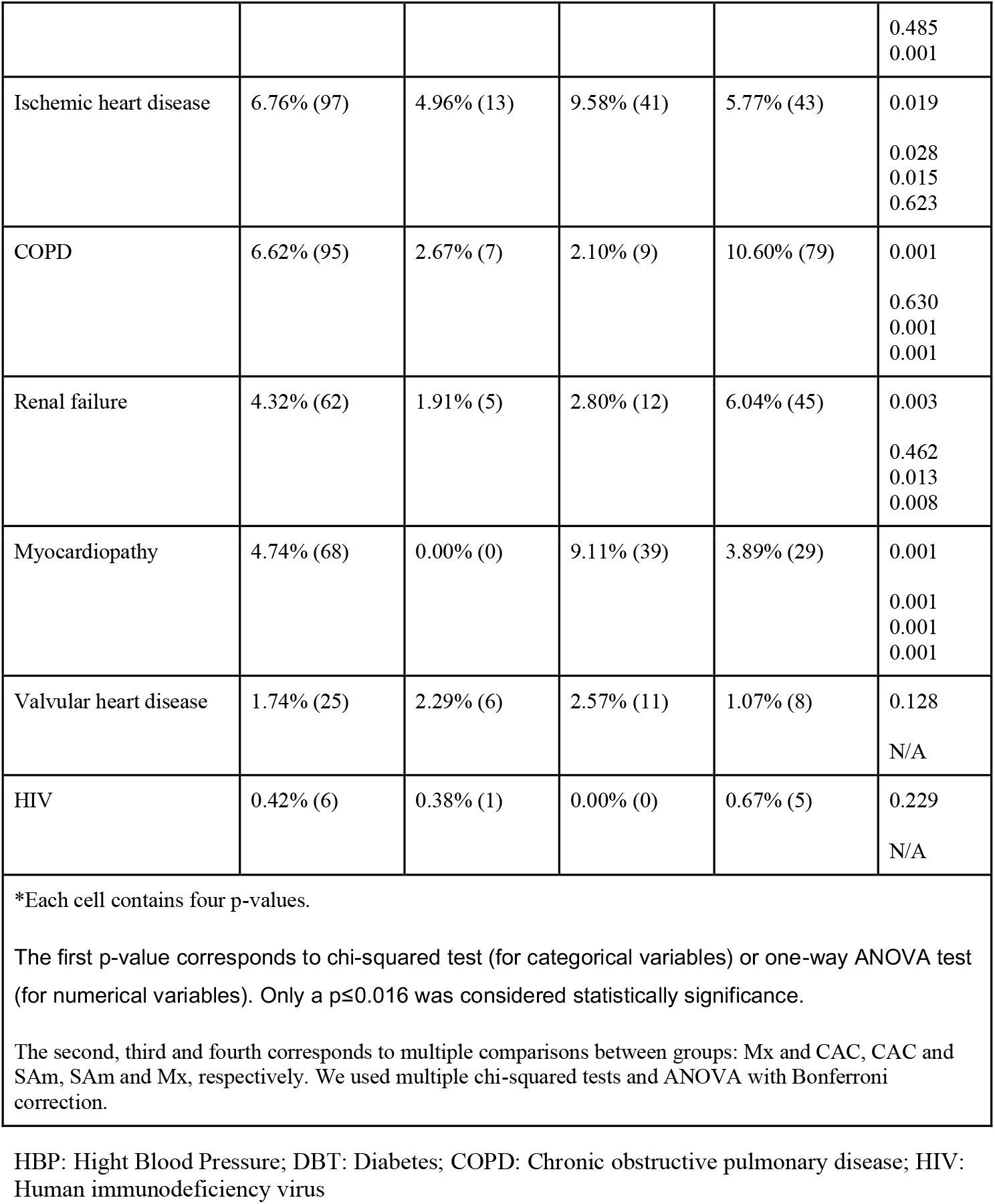
Demographic variables and baseline Comorbidities

### Cardiopulmonary Imaging Utilization

cCT (61%) and cX-ray (46%) were the most common cardiopulmonary imaging performed in our cohort; cEcho was carried out in 18% and LUS in 7% of patients. The use of imaging modalities was different across regions: cCT was more frequent in Mx and SAm, and cX-ray was significantly more common in CAC. cEcho was almost the same among regions with a small predominance in Mx. LUS was significantly more common in Mx. Brazil had the highest use of both modalities if we analyze based on country. ECG use was limited (24%) due to the biosafety measures implemented during COVID-19. The lowest use was in SAm (except Brazil). We could also consider an underreporting of ECG due to a lack of digitization (Table 2). cMRI was performed in only one patient in the cohort and was not taken into account for the statistical analysis.

**Table 2:**
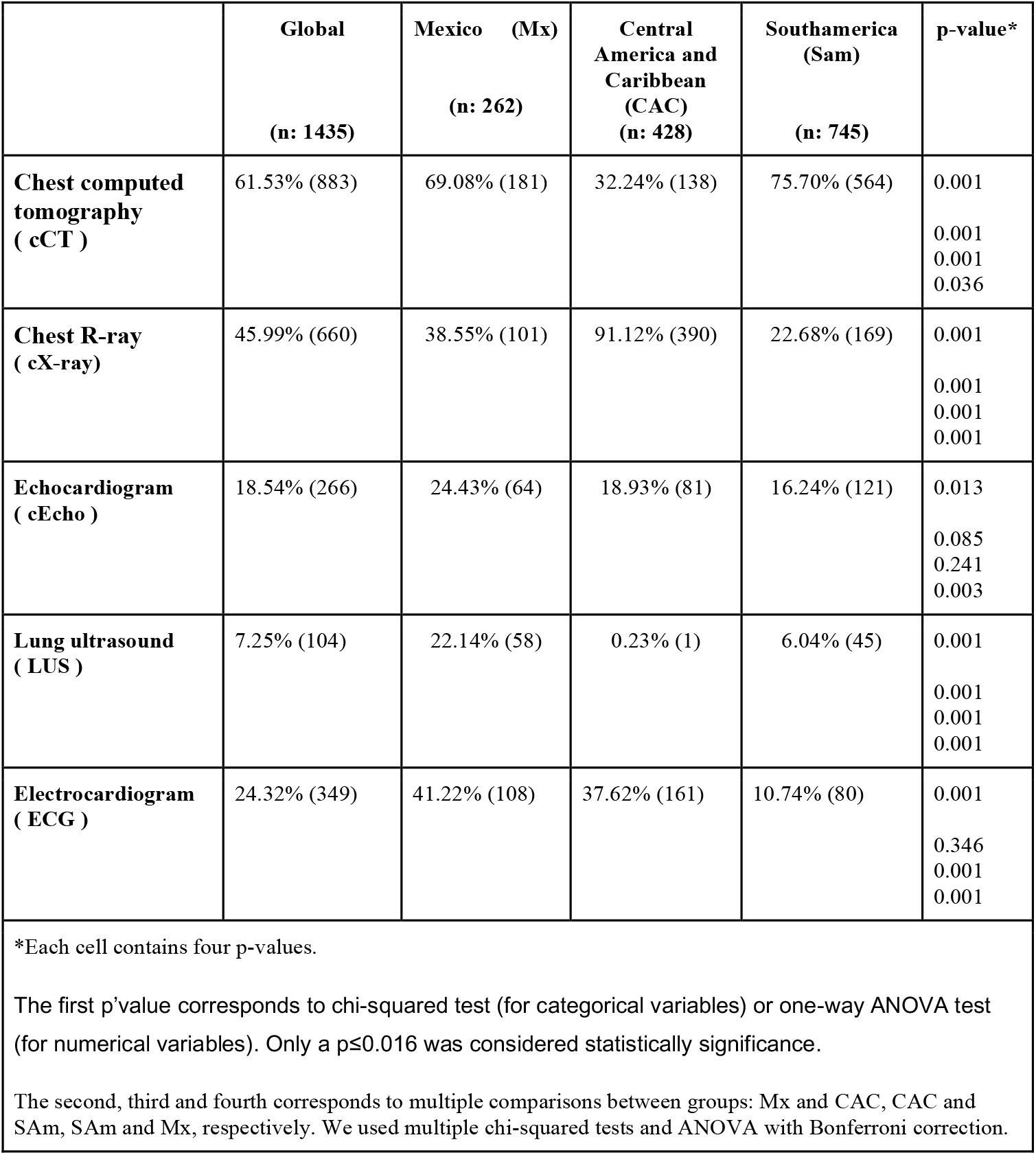
Image modalities used

A multivariate analysis of the use of images was carried out considering confounding factors such as: age, sex, hypertension, overweight / obesity, diabetes, renal failure, heart failure, hospital stay, ICU stay, mechanical ventilation and death (Table 3). The calculated adjusted ORs do not appear to add significant variation or impact.

**Table 3:** Images used and confounders: multivariate analysis

|  | Zone 1 (Mx) |  | Zone 2 (CAC) |  |  |  |
| --- | --- | --- | --- | --- | --- | --- |
|  | n: 262 | crude OR | n: 428 | crude OR<br>(95%CI) | adjusted OR*<br>(95%CI) | n: 745 |
| Chest computed tomography ( cCT ) | 69.08% (181) | Baseline | 32.24% (138) | 0.21<br>(0.15-0.29) | 0.19<br>(0.13-.27) | 75.70% (564) |
| Chest R-ray ( cX-ray) | 38.55% (101) | Baseline | 91.12% (390) | 16.36 (10.79-24.79) | 18.09<br>(11.67-28.03) | 22.68% (169) |
| Electrocardiogram ( ECG ) | 41.22% (108) | Baseline | 37.62% (161) | 0.85<br>(0.62-1.17) | 0.68<br>(0.48-0.96) | 10.74% (80) |
| Echocardiogram ( cEcho ) | 24.43% (64) | Baseline | 18.93% (81) | 0.72<br>(0.49-1.04) | 0.82<br>(0.55-1.23) | 16.24% (121) |
| Lung ultrasound ( LUS ) | 22.14% (58) | Baseline | 0.23% (1) | 0.01<br>(0.01-0.05) | 0.01<br>(0.01-.06) | 6.04% (45) |

\* Confounders: Age, Sex, hypertension, overweight/obesity, diabetes, renal failure, cardiac failure, hospital stay, ICU, MV, death

### Findings

The imaging findings and their regional distribution are shown in Table 4. The most frequent patterns of the cX-ray were peripheral, basal, and ground glass infiltrates with a significant prevalence of abnormalities in Mx. The ground glass appearance of peripheral or subpleural infiltrates on cCT was found in 89% of cases; the most severe subtypes (infiltrates > 50 %) were significantly higher in Mx (Figure III). The left ventricular ejection fraction (LVEF) was calculated in all cEcho performed (266 patients) with a mean value of 57% being almost the same in the three regions.

**Table 4:**
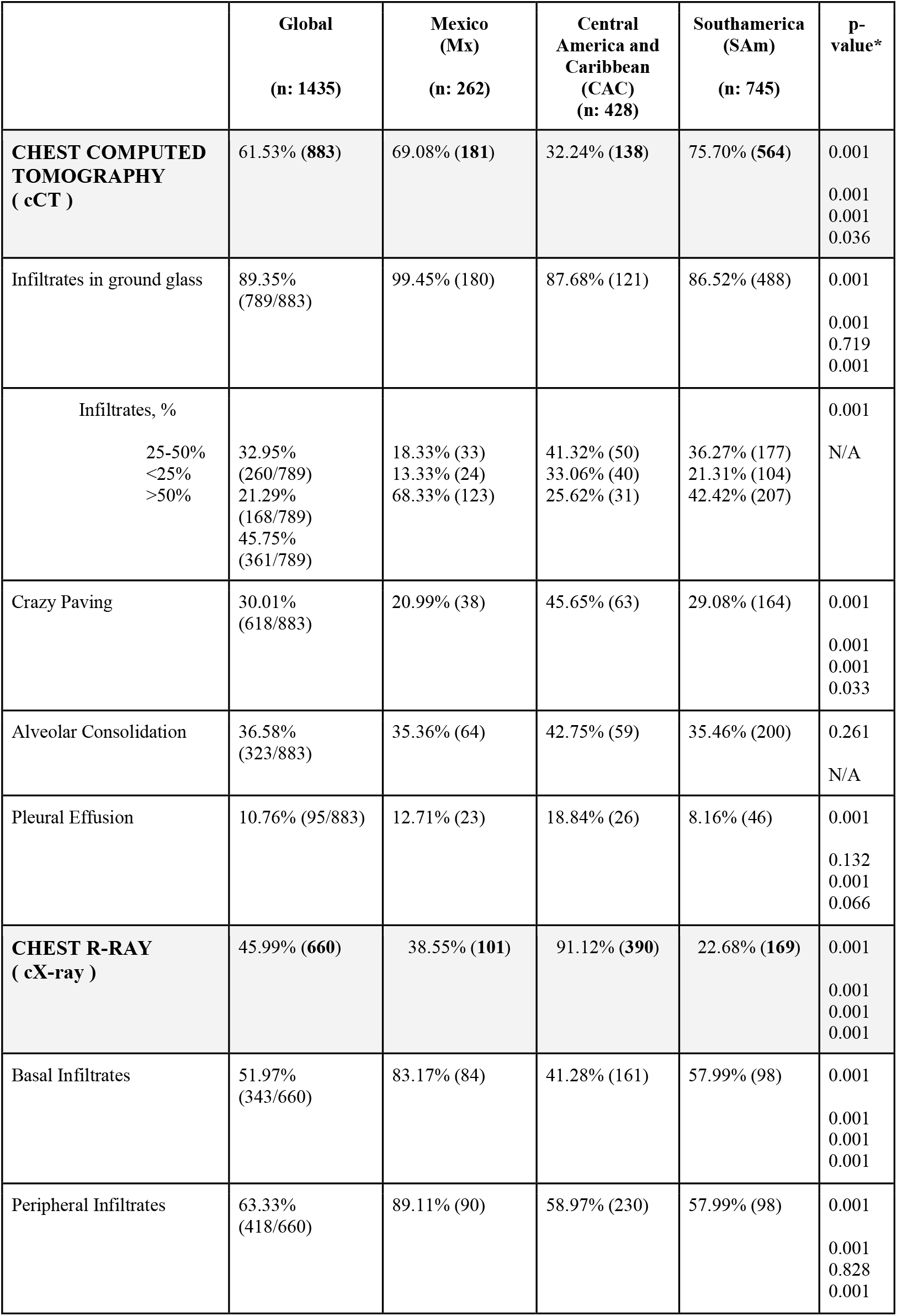
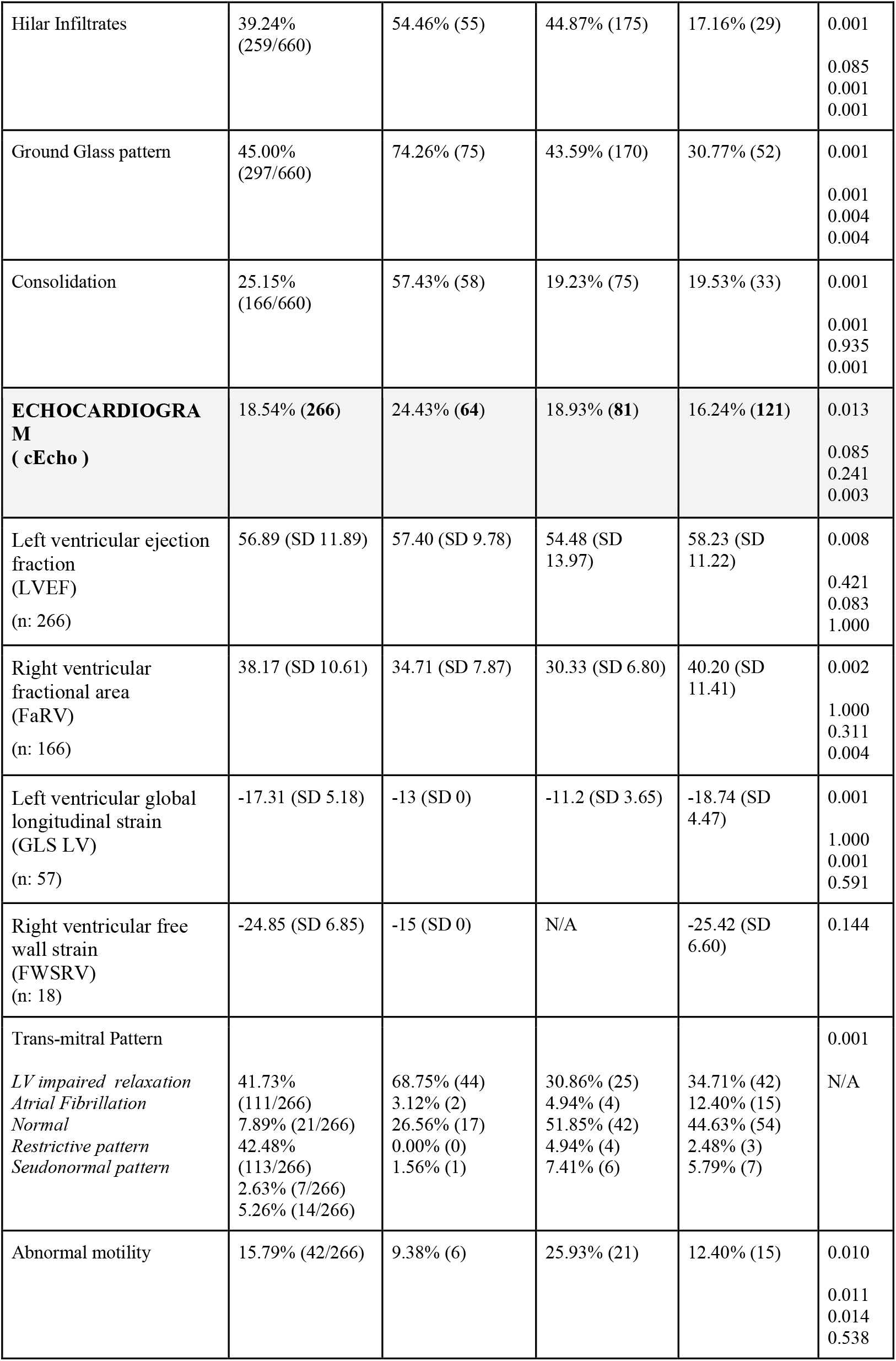
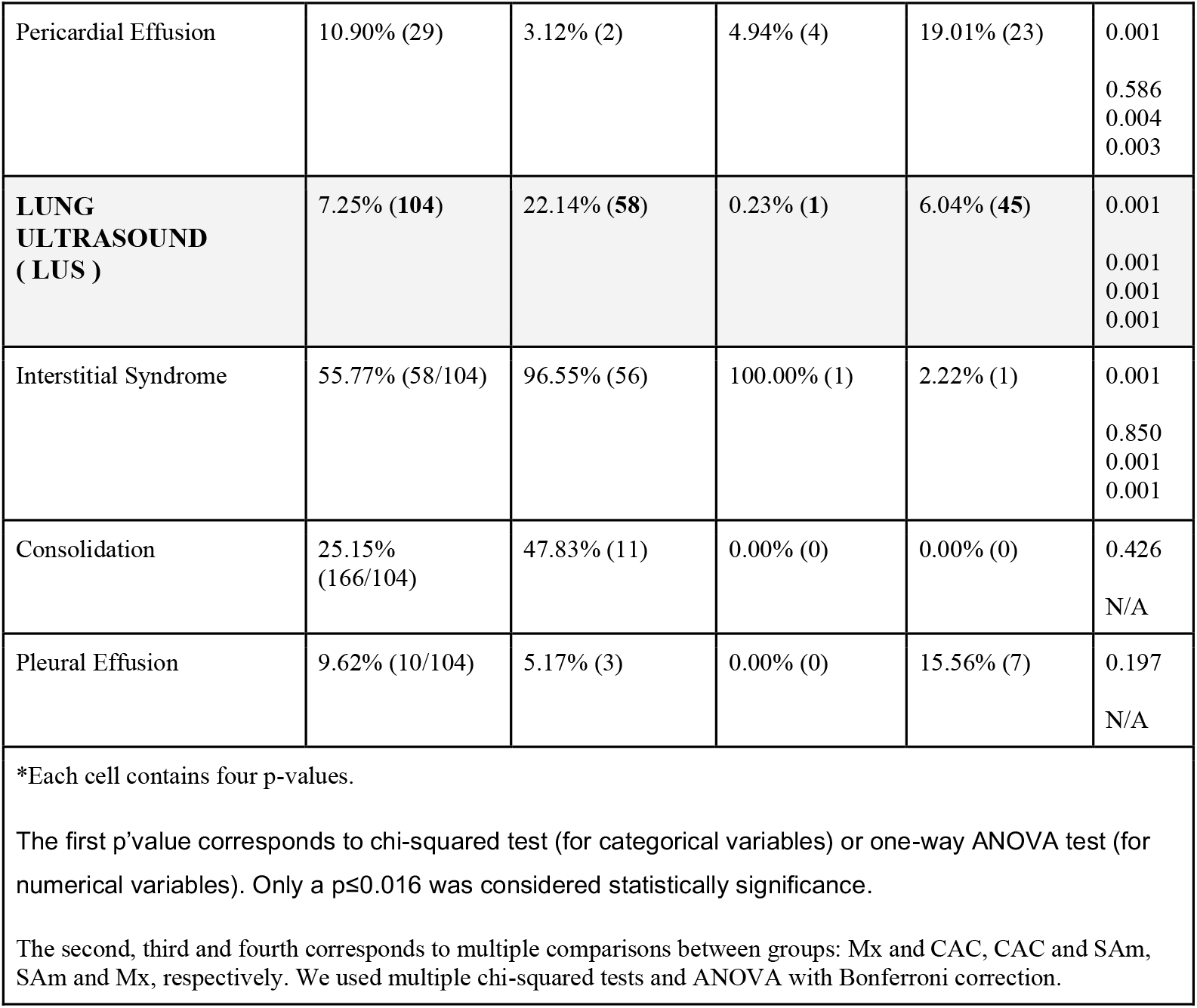
Image Patterns Global and Regions

**Figure III:**
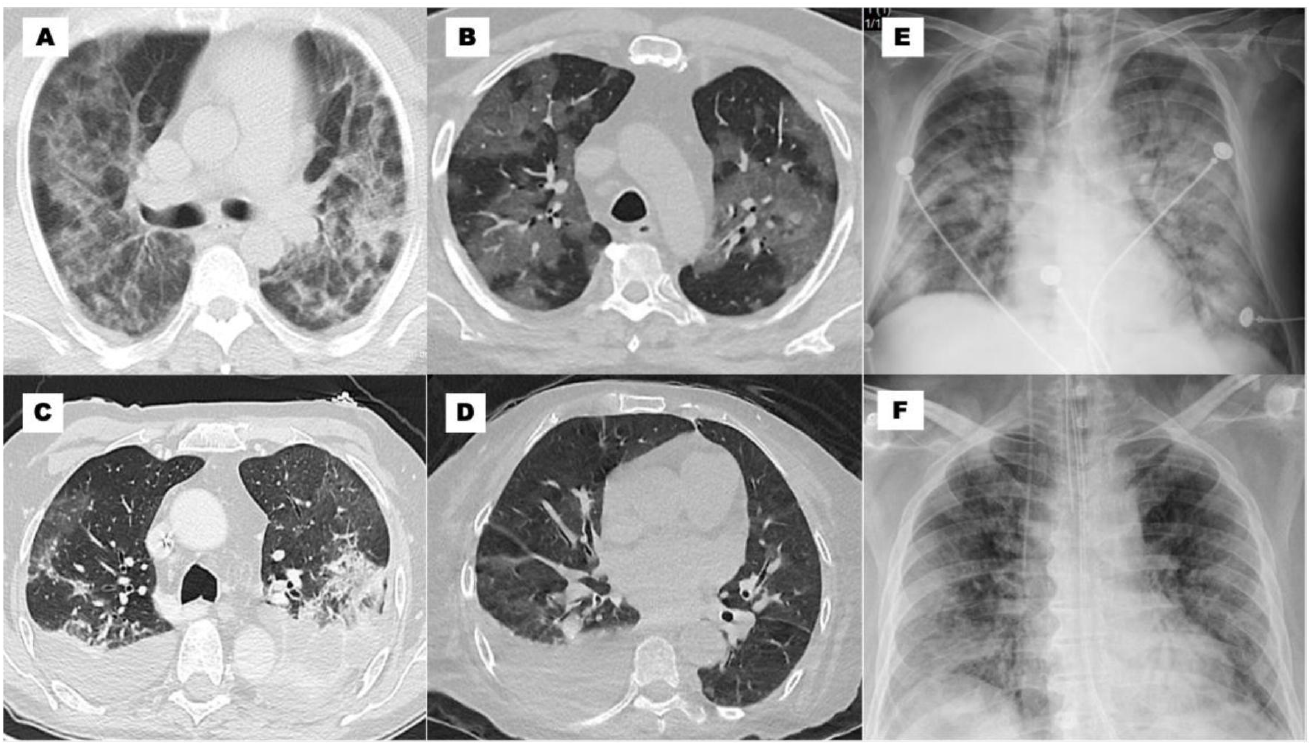
Image Patterns. **A: cTC:** infiltrates in ground glass > 50%, crazy paving and alveolar consolidation bilateral; **B: cTC:** infiltrates in ground glass > 50% bilateral; **C: cTC:** infiltrates in ground glass 25 - 50% and pleural effusion bilateral; **D: cTC:** infiltrates in ground glass < 25% and pleural effusion unilateral; **E: cX-ray:** peripheral, basal and hilar infiltrates, ground glass pattern and consolidation; **F: cX-ray:** basal, and peripheral infiltrates, and ground glass pattern.

The right ventricular fractional area (FaRV) was performed in almost 62% of the cEcho (166 patients) with a mean value of 38% and no differences between regions. Strain echocardiography was performed only in Mx and SAm. The left ventricular global longitudinal strain (GLSLV) was performed in 21% of cEcho (57 patients) with a median of -17 (−7 to -26) and lower in Mx (−13). The right ventricular free wall strain (FWSRV) was achieved in 7% of cEcho (18 patients) with a median value of - 25 (−7 to – 40); it was also lower in Mx (−15). The trans-mitral/tissue Doppler patterns most frequently were majority normal or impaired relaxation (42% each). The development of abnormal regional wall motility was predominant in CAC. Pericardial effusions was found in 11% of the patients—the majority in SAm. The most frequent patterns in LUS were the interstitial syndrome (predominant in Mx and CAC) and consolidation (predominant in Mx). Logistic regression showed associations of imaging with comorbidities, complications, and evolution with an estimation of crude OR and their 95%CI shown in Table 5.

**Table 5:**
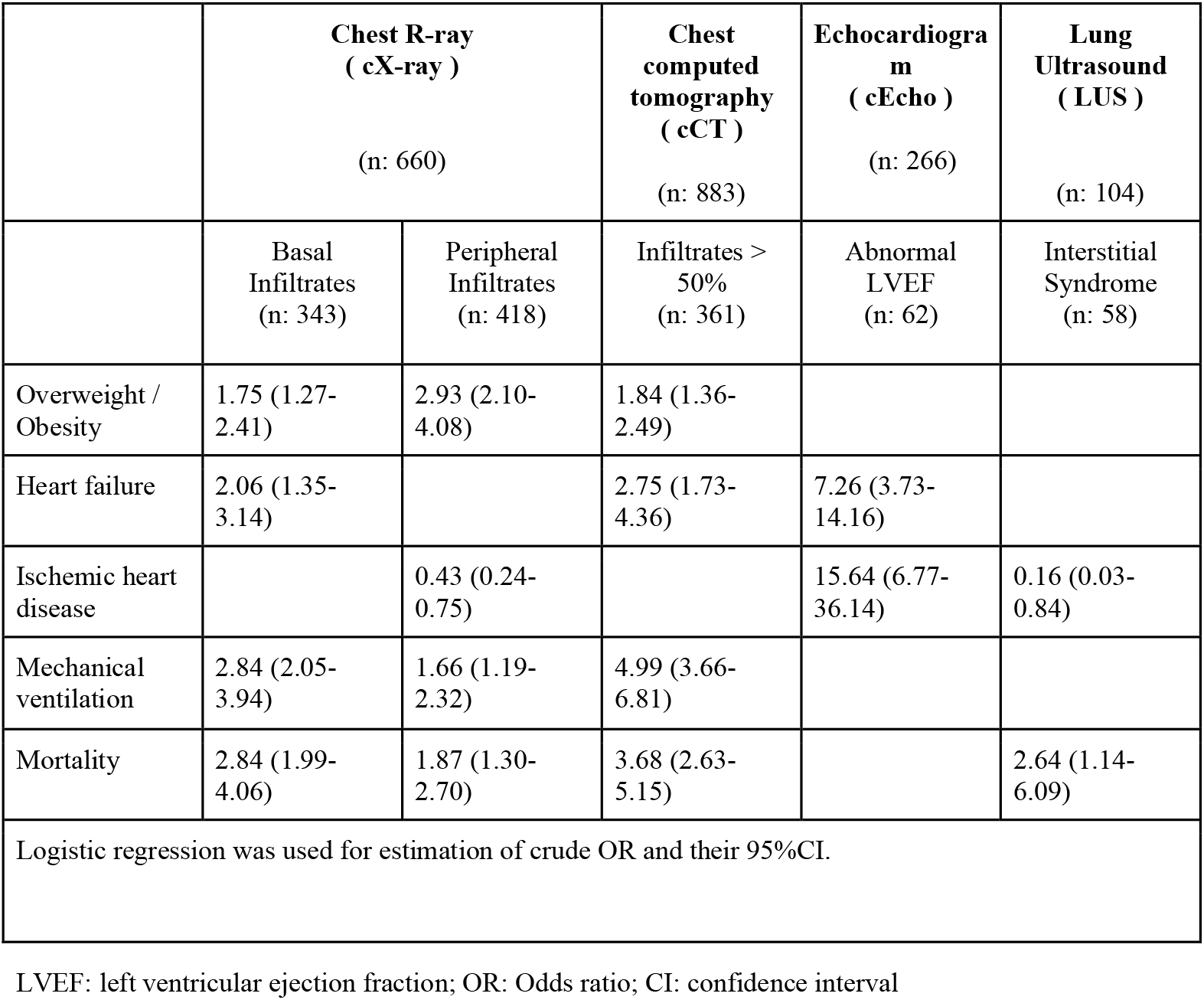
Modalities imaging logistic regression associated with comorbidities, complications and evolution

The most frequently finding in ECG was an arrythmia especially in SAm.

### Other parameters

The most prevalent complication was renal failure (20%) with a significant predominance in Mx and SAm regions. Heart failure developed in 13% (191 patients) of the cohort with a predominance in Mx and CAC; left ventricular failure occurred in 46%, right ventricular failure in 20%, and biventricular failure in 34% of cases.

Lung thromboembolism was significantly higher in Mx while acute myocardial infarction was predominant in CAC (Table 6).

**Table 6:**
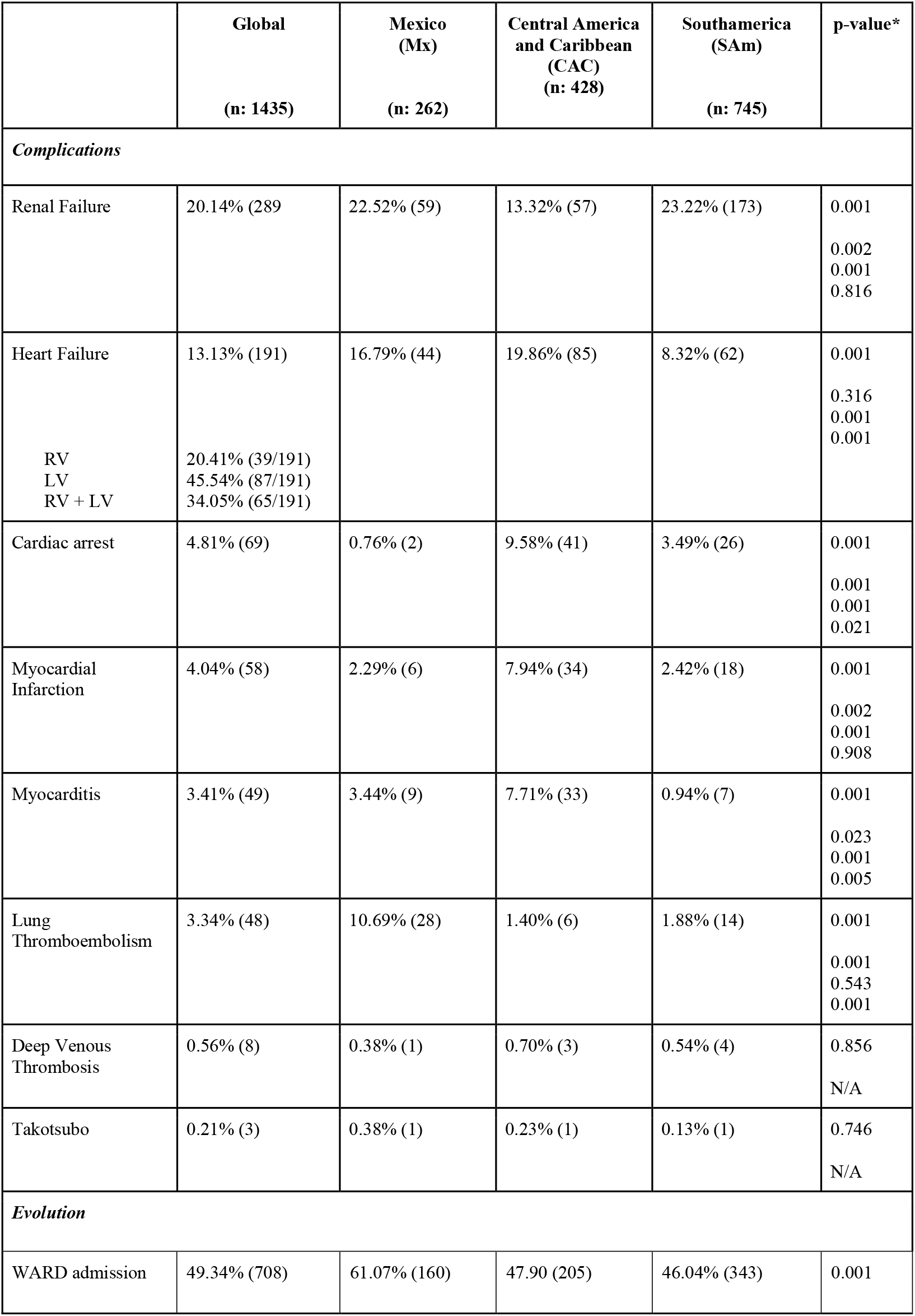
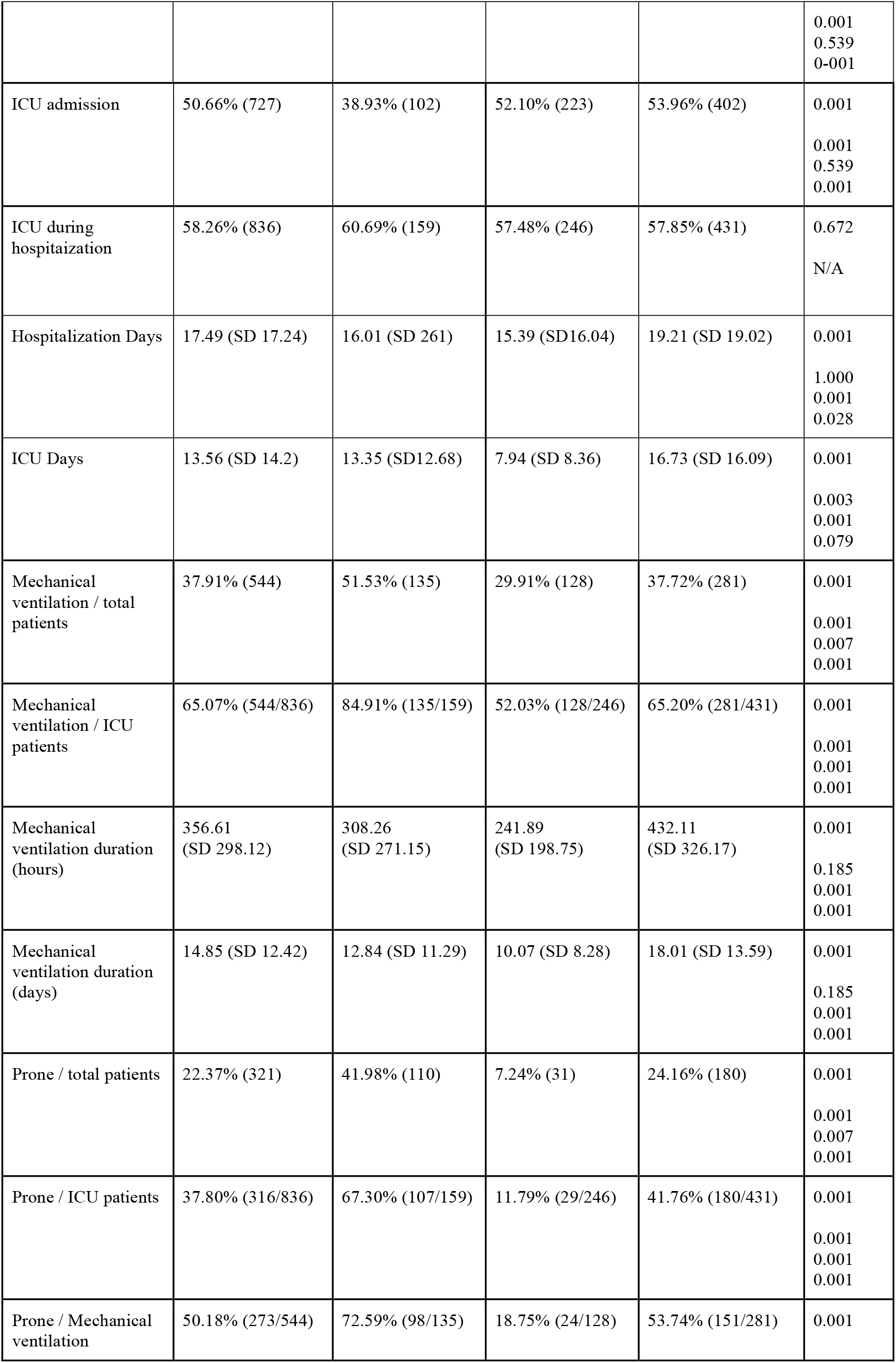
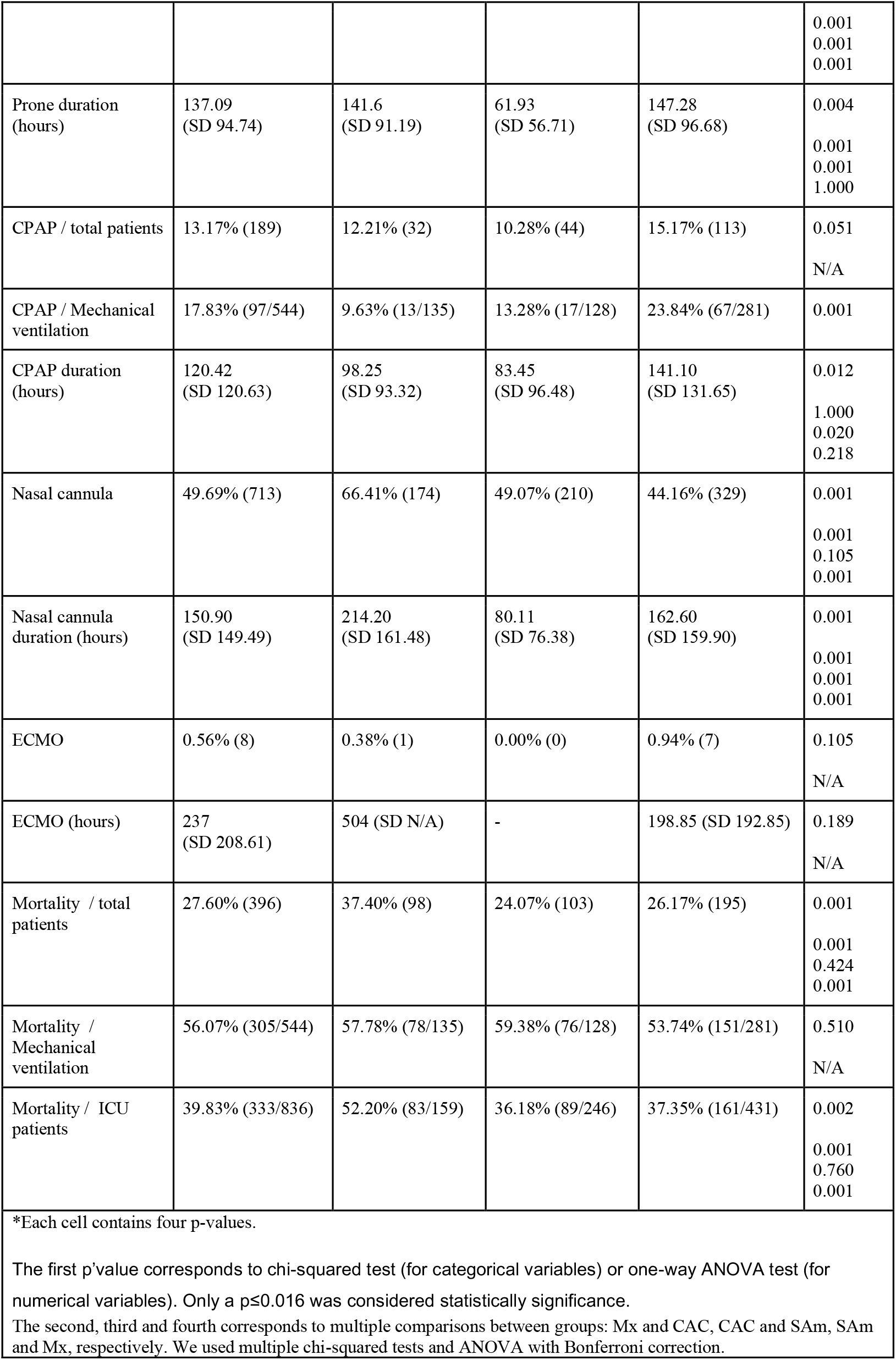
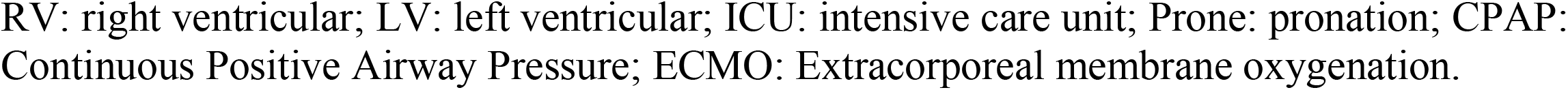
Complications and Evolution

Modalities of respiratory support had significant differences across regions: Overall, mechanical ventilation (MV) and pronation predominated in Mx ICU patients (85%-67% respectively), although the mechanical ventilation duration (hours and days) were higher in SAm. CPAP in mechanical ventilated patients was more common in SAm; ECMO was rarely employed (Table 6)

The overall in-hospital mortality was 28% (CI:25.2-29.9) and was significantly higher in 65 years with no differences between gender; this is nearly similar to that published in patients from the United Kingdom (19) and slightly higher than that of Iran (20). The higher regional mortality was in Mx (37%) with no significant differences between CAC and SAm. Rather, if we analyze for each country, then Perú and Argentina had the highest mortality (40% and 39% respectively). The ICU mortality was 40% and was significantly higher in México (52%). Mortality in patients with mechanical ventilation was 56% with no differences between regions (Table 6). The risk factors associated with high mortality were HBP, DBT, overweight/obesity, as well as hospitalization location and the requirement of MV.

## Discussion

The SARS-Cov-2 pandemic has had a global influence, but little is known about the impact on LATAM. The respiratory system is dramatically impacted by COVID-19, and this explains the necessary use of cX-ray and cCT overshadowing the use of cEcho and LUS. In this multicenter study, we describe the different modalities of imaging used and their findings in the management of patients affected during the early stages of the COVID-19 disease in LATAM. The three regions had expected as well as unexpected results.

The diagnosis and follow-up of pulmonary involvement was carried out using lung imaging (chest X-ray or cCT). The procedures optimized the technical and human resources available in light of the personnel protection measures. Only in doubtful or complex cases were both techniques used in the same patient (18%). This could partly explain the difference in the use of these modalities between the three regions. The findings of basal and peripheral infiltrates on chest X-ray and of ground glass infiltrates > 50% in cCT were correlated with the presence of overweight/obesity, greater occurrence of heart failure, need for mechanical ventilation, and higher mortality (Table 5).

According to SISIAC’s recommendations, the use of cEcho during the COVID-19 pandemic in 2020 was limited to patients with hemodynamic instability, new heart failure or ischemic heart disease, complex arrhythmias, or a high suspicion of endocarditis associated with coronavirus. The higher prevalence of heart failure, myocardial infarction, and pulmonary thromboembolism in Mx and CAC could probably explain the slightly higher use of cEcho in these regions. The findings on this technique showed that the analyzed cohort had a normal mean ejection fraction and diastolic function on admission. The development of a reduced ejection fraction during hospitalization was correlated with the presence of heart failure and ischemic heart disease (Table 5).

Uncertainties about its usefulness and the lack of practice of LUS in patients with pulmonary and cardiac pathology in most LATAM countries contributed to the limit use of this technique except in Mexico and Brazil. Of note, published studies (21) that used LUS in COVID-19 patients were oriented to usefulness and results, and remains unclear what the real rate of LUS use was. Interstitial infiltrates were correlated with higher mortality (Table 5).

### Regarding regions

We hypothesize that the higher mortality in Mx could be explained by a combination of these findings: a higher prevalence of cX-ray infiltrates; cCT infiltrates >50% in more than two-thirds of the subjects; high incidence of interstitial syndrome in LUS; highest ICU stay in days; highest proportion of total patients admitted to the ICU with mechanical ventilation and pronation use (more severe ICU patients); and a higher prevalence of pulmonary thromboembolism, heart failure, and renal failure. Furthermore, the high incidence of comorbidities such as obesity, diabetes, and tobacco use suggest a high-risk cohort.

The CAC and SAm regions had fewer numbers of complications; there was less prevalence of abnormal cX-ray findings, and a higher rate of moderate cCT infiltrates (25-50%). These regions had a lower proportion of total patients admitted to the ICU with mechanical ventilation and pronation use (less severe ICU patients); these could probably identify a moderate risk cohort. Interestingly, we found that the CAC region had the highest rate of Hypertension and the highest proportion of regional wall motion abnormalities on cEcho while SAm had the highest rate of arrhythmias as well as the highest prevalence of pericardial effusions. We hypothesize that the combination of these factors could probably explain the similar mortality between both regions (24% and 26%, respectively).

Remarkably, the cardiopulmonary images used, and their findings had a great impact on the diagnosis and prognosis of COVID-19 disease as well as on the mechanical ventilation and pronation necessary to treat it. The addition of comorbidities and complications could explain the different severity rates in these patients.

## Data Availability

All data produced in the present study are available upon reasonable request to the authors

## Abbreviations

BMI: Body Mass Index
CAC: Central America and Caribbean region
cCT: chest computed tomography
cEcho: echocardiogram
cMRI: cardiac magnetic resonance imaging
COPD: Chronic obstructive pulmonary disease
cX-ray: chest R-ray
DBT: diabetes
ECG: electrocardiogram
FaRV: right ventricular fractional area
FWSRV: right ventricular free wall strain
GLSLV: left ventricular global longitudinal strain
HBP: high blood pressure
ICU: intensive care unit
IRB: institutional Review Board
ISR: imaging services resources
LUS: lung ultrasound
LVEF: left ventricular ejection fraction
MV: mechanical ventilation
Mx: Mexican region
SAm: South America region

## Strengths

To the best of our knowledge, this is the first international, multicenter study in LATAM to characterize cardiopulmonary imaging use and their findings in patients hospitalized with COVID-19.

## Limitations

The cohort might not be representative of all COVID-19 hospitalizations in each country. In addition, data were collected during the first pandemic wave and therefore findings may not reflect changes that may have occurred later.

## Future Directions

Subsequently, imaging societies may consider developing risk scoring protocols based on the findings of the images performed.

## Conclusion

Patients hospitalized with COVID-19 had differences in the images used in the three LATAM regions. These could be explained by clinical needs, personnel protection measures and/or hospitalization location. cCT and cX-ray were the most frequently performed modalities, and cEcho was employed in just special clinical situations. Abnormal findings had a great impact on diagnosis and prognosis, on the use of mechanical ventilation, the necessary pronation, and overall mortality.

